# Early assessment of knowledge, attitudes, anxiety and behavioral adaptations of Connecticut residents to COVID-19

**DOI:** 10.1101/2020.05.18.20082073

**Authors:** Toan Ha, Stephen Schensul, Judy Lewis, Stacey Brown

## Abstract

**Objectives:** To assess knowledge, attitudes, anxiety and behavioral adaptations to COVID-19.

**Methods:** A cross-sectional study was conducted among non-healthcare-related participants after a stay-at-home directive was implemented. Multivariate logistic regression analysis was conducted to identify factors associated with anxiety, perceived seriousness of COVID-19 and loneliness.

**Results:** A total of 464 participants responded to the survey. Most participants recognized cough, shortness of breath and fever as primary symptoms of COVID-19. Nearly 50% reported high levels of anxiety to COVID-19 and 48% reported being loneliness during the social isolation. Higher level of COVID-19 knowledge was associate with higher levels with anxiety. Being married had 1.79 times higher levels of anxiety about COVID-19. Women were less likely to report loneliness than men. Older age was associated with taking the pandemic seriously, and was also associated with loneliness during the social isolation.

**Conclusions:** It is crucial for the public health authorities not only provide accurate and scientific information about the COVID-19 promoting protective behavior changes but also to minimize anxiety through supportive messages and recommendations for positive coping strategies and timely offering mental health counselling services for those in need.

## Introduction

Covid-19 is an infectious respiratory disease which is currently understood to spread from person-to-person through respiratory droplets expelled during coughs or sneezes by an infected person.^1^ The outbreak of COVID-19, first occurring in Wuhan, Hubei Province, China^2^ has spread rapidly around the world.^3^ The World Health Organization (WHO) declared Covid-19 a pandemic on March 11, 2020.^4^ The first case of COVID-19 in the in the United States was reported on January 21, 2020.^5^ As of April 24, 2020, a total of 871,285 cases and 50,066 deaths were reported in the US ^6^ and the disease is now present in every state in the U.S.^5^

As of April 24, 2020, Connecticut reported 23,100 cases including 1639 deaths.^7^ In response to the COVID-19 widespread outbreak in the State, the Connecticut governor issued several executive orders including the stringent “Stay safe and stay home” initiative declaring a public health and civil preparedness emergency. This declaration mandated the reduction of social and recreational gatherings, closure of large, indoor shopping malls, cancellation of classes at all schools and directed all non-essential workers to work from home.^8^

The response to the pandemic has involved health care system administrators, clinicians, laboratory researchers, and government officials, addressing a wide range of needs from personal protection equipment (PPE) for health care providers, ventilators for patients and intensive care unit (ICU) bed availability to testing of treatment approaches, development and distribution of diagnostic tests to the search for an effective vaccine. At the same time, governmental officials and media have been communicating information on the status of the pandemic globally and locally, conveying epidemiological reports, guidelines for preventing infection and providing information on the capacity of the health care system to meet the demands of patients with COVID-19.

The current COVID-19 as well as other viral epidemics in the last two decades including H1N1, MERS, Ebola, H7N9 pose difficult challenges for communication to the public.^9^ While information to the public is critical, it is also essential that the information be factual and understandable. The scientific uncertainty and ambiguity associated with novel viruses can create a lack of trust in official guidelines, unclear appreciation of risk or fatalistic perceptions. Political infighting producing competing policies and actions further complicates public understanding. With no antiviral treatment or vaccine recommended for COVID-19 at the moment, the uptake of preventive measures to control COVID-19 infection is critical.^10^

As a result, it is essential that we assess the impact of official and unofficial communication concerning COVID-19 so that we can discover problems, improve messaging, address gaps and identify subpopulations where information is not addressing needs and challenges.^11^ The purpose of this paper is to assess knowledge, attitudes and behavioral adaptations to COVID-19 among Connecticut residents. Findings of this study will contribute to the literature on public knowledge, attitudes and behavioral adaptations, and assist in developing more effective public health messages on COVID-19.

Faculty in the Department are epidemiologists, social scientists and lawyers, involved in research on communicable and non-communicable diseases, medical ethics, and environmental and occupational health. The faculty direct and teach in public health programs at the undergraduate and graduate (MPH and PhD) levels. While the basic science and clinical departments were contributing their expertise to global, national and local COVID-19 challenges, the Department recognized that there was little understanding of the ways in which the public in Connecticut were responding to the flood of COVID-19 messaging.

## Methods

In response to the lack of information about public response, the faculty with student input developed a survey instrument that would assess the public’s knowledge, attitudes and behavioral adaptations. With little time, no funds, only virtual meetings and the need for data in “real-time,” a collective decision was made that each faculty and student of the Department would recruit three to five contacts from the public (non-healthcare related) to respond to the survey. While this recruitment procedure was not random, this approach would provide rapid feedback as well as test the validity and reliability of the survey items. The survey was rolled out on March 23, 2020, one week after a stringent stay-at-home directive was implemented in Connecticut and closed to recruitment on March 29, 2020.

### Sample

The inclusion criteria involved participants aged 21 or over, in the social network of the faculty, and students and residing in Connecticut, but outside of their family or kin. Participants were contacted (e-mail, phone, skype or other means) by the faculty and students, and the telephone consent script was read to them. If consent was obtained, participants could choose a hyperlink for self-administration of the survey online or faculty or students could administer the survey as an interview over the phone or other means of communication and record their answers online. The study was approved by the UConn Health Institutional Review Board.

### Measures

Demographic variables included age, education, gender, marital status, employment, and ethnicity.

### Perceived risk, seriousness of COVID-19, anxiety and loneliness

Respondents were asked questions regarding self-perception of the seriousness of COVID-19 for the country, the state, community, family and themselves with a 5-point Likert scale ranging from “Not likely” to “Very likely”. One question asked the perceived risk of acquiring disease with a 5-point Likert scale ranging from “Not likely” to “Very likely”. One question asked the anxiety level associated with COVID-19 using a 5-point Likert scale ranging from “Very low” to “Very high”. One Yes/No question about loneliness faced during the social isolation.

### Knowledge of COVID-19 and Preventive behavioral responses

Respondents’ knowledge of COVID-19 was assessed with a series of nine True/False questions. Preventive behavioral responses to COVID-19 were assessed with a series of 12 Yes/No questions. Health seeking behaviors when having coronavirus-like symptoms were assessed using a series of 16 True/False questions. Participants were also asked one question about how long they stockpile of food and household items during the social isolation ranging from one to two days to three to four weeks.

### Sources of information and trustworthy of communication about COVID-19

A series of seven Yes/No questions asking each source of information where participants received information about COVID-19. Trust of the communications concerning COVID-19 were assessed using a series of 13 questions with a 5-point Likert scale ranging from “Not trust” to “Highest trust”.

The questionnaire was developed using Qualtrics software. ^12^ The draft survey instrument was circulated to the Department faculty to assess its understandability and validity and revised before it was distributed to the study population.

### Data analysis

The data was downloaded into SPSS 26.0 for analysis. Descriptive statistics were used to assess the socio-demographic characteristics, risk perception, knowledge, sources of information and behavioral changes. One-way analysis of variance (ANOVA) and Chi-square test compared means for continuous and nominal outcomes as appropriate. Multivariate logistic regression analysis was conducted to identify factors associated with anxiety, seriousness of COVID-19 and loneliness controlling for age, gender, education, marriage, employment and ethnicity. A p value of < .05 was considered to indicate statistical significance.

## Results

A total of 464 participants responded to the survey. The mean age was 41.0. Over three fourths (76.5%) of participants had a bachelor’s degree or higher. Sociodemographic characteristics are shown in Table 1.

**Table 1.**
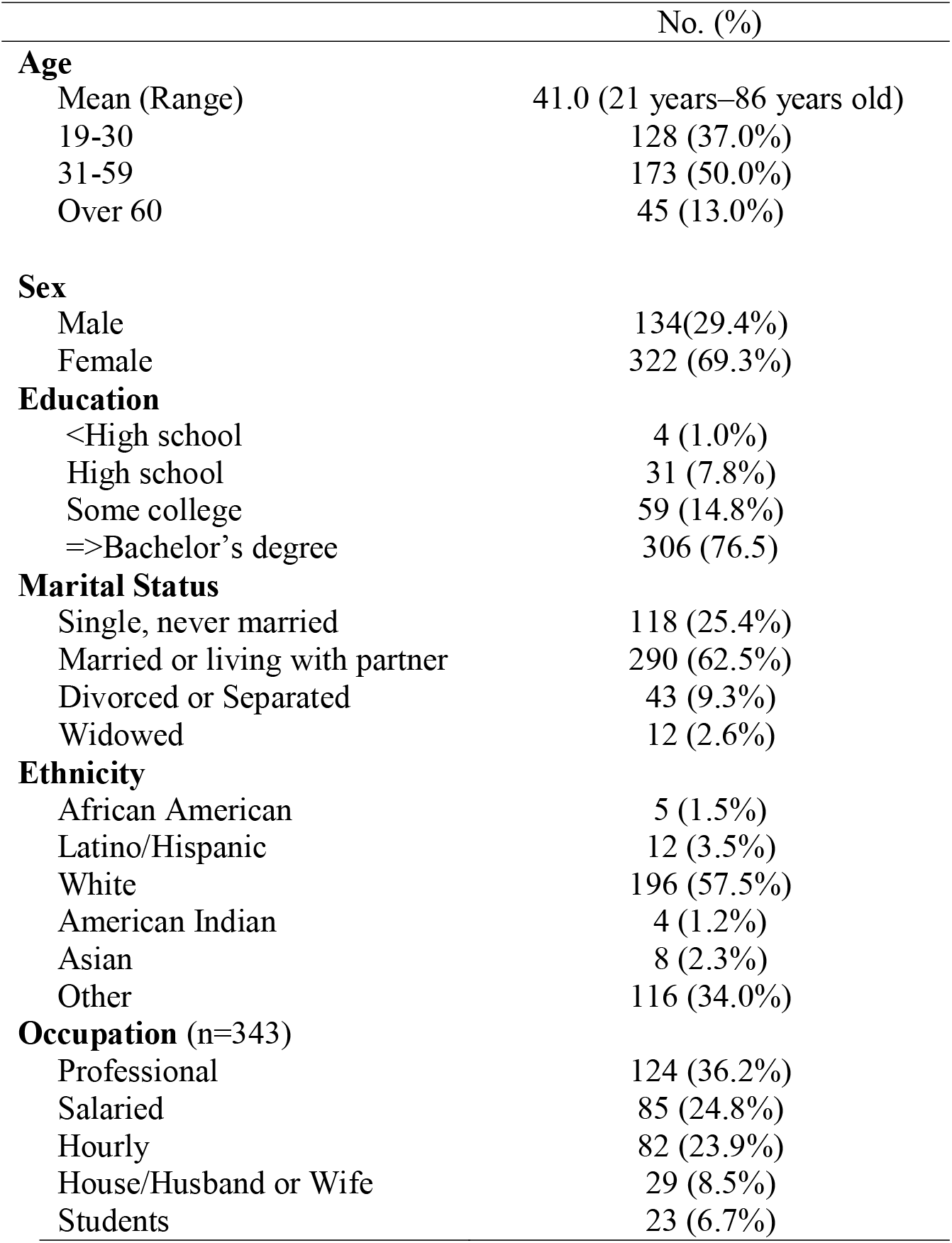
Characteristics of participants (n=464)

|  | No. (%) |
| --- | --- |
| **Age** |  |
| Mean (Range) | 41.0 (21 years–86 years old) |
| 19-30 | 128 (37.0%) |
| 31-59 | 173 (50.0%) |
| Over 60 | 45 (13.0%) |
| **Sex** |  |
| Male | 134(29.4%) |
| Female | 322 (69.3%) |
| **Education** |  |
| <High school | 4 (1.0%) |
| High school | 31 (7.8%) |
| Some college | 59 (14.8%) |
| =>Bachelor's degree | 306 (76.5%) |
| **Marital Status** |  |
| Single, never married | 118 (25.4%) |
| Married or living with partner | 290 (62.5%) |
| Divorced or Separated | 43 (9.3%) |
| Widowed | 12 (2.6%) |
| **Ethnicity** |  |
| African American | 5 (1.5%) |
| Latino/Hispanic | 12 (3.5%) |
| White | 196 (57.5%) |
| American Indian | 4 (1.2%) |
| Asian | 8 (2.3%) |
| Other | 116 (34.0%) |
| **Occupation (n=343)** |  |
| Professional | 124 (36.2%) |
| Salaried | 85 (24.8%) |
| Hourly | 82 (23.9%) |
| House/Husband or Wife | 29 (8.5%) |
| Students | 23 (6.7%) |

### Sources of COVID-19 information

Most of the participants received information through electronic media and TV (85.5%), social media (81.3%), family members (69.0%) and friends (68.7%) (Supplement Table A). Males reported social media more often than females (X^2^=6.17; p<0.05). Those with less education reported a higher level of TV as a source of information [F1, 422) =4.04, *p* =.045]. There were no significant differences in use of social media based on education level [F1, 422) =2.10, *p* = .147]. In terms of trustworthiness of sources of information, the President/Vice-President and Congress had the lowest ratings (12.8% and 12.7%), while government agencies were more highly rated including Dr. Anthony Fauci of the National Institute of Health (NIH) (82.3%), the Center for Prevention and Control (CDC) (84.9%) and the Connecticut State Health Department (79.4%) (Figure 1).

**Figure 1.**
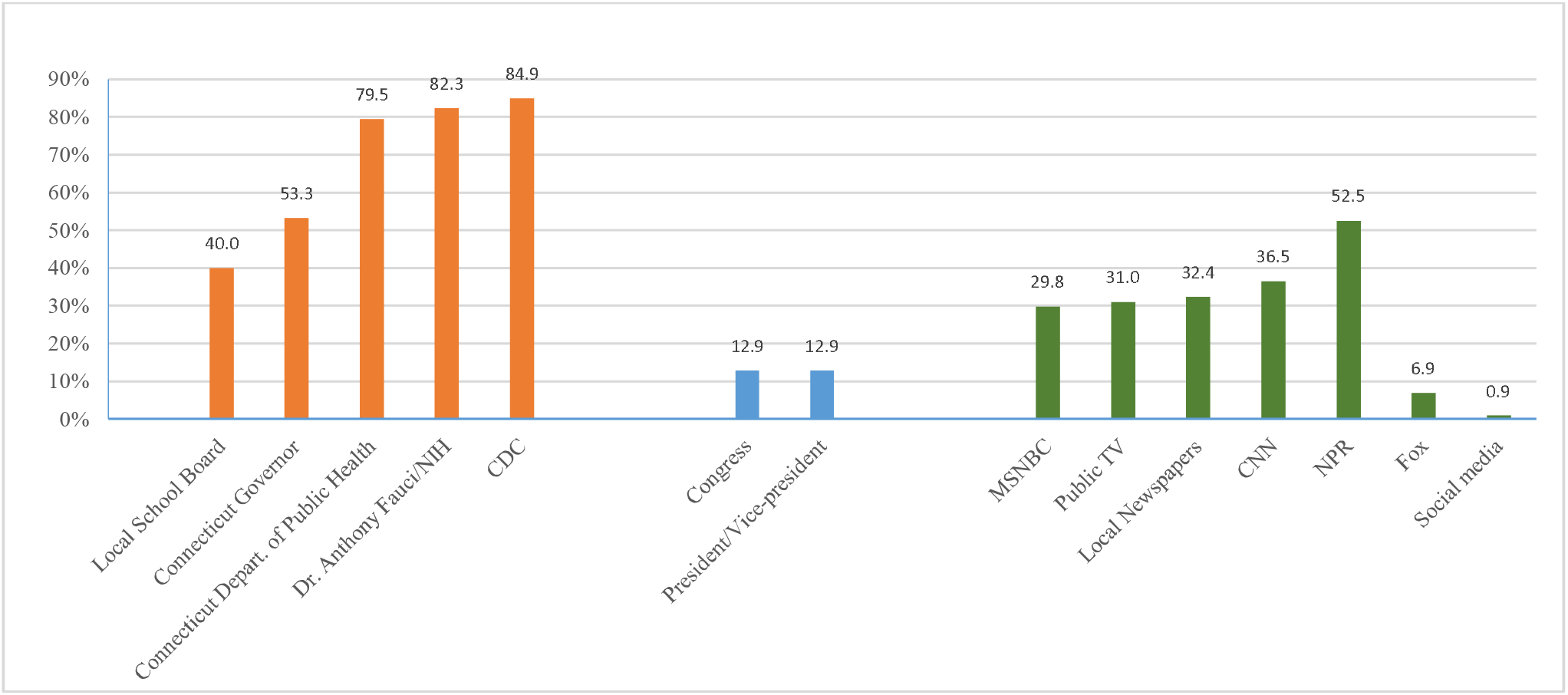
Trust of the communications concerning COVID-19

### Perceived risk, seriousness of COVID-19, anxiety and loneliness during the social isolation

When asked how likely is it that you will contract the coronavirus, only 35.1% stated that they were likely or very likely to contract the coronavirus. In response to the question about perceived seriousness of the pandemic, nearly 38% of participants did not consider COVID-19 serious for themselves (Figure 2). In response to the question of rating your current anxiety level related to the COVID-19 pandemic, 222 (47.8%) participants stated being high and very high. Fourth eight percent participants reported being loneliness during the shutdown and social isolation.

**Figure 2.**
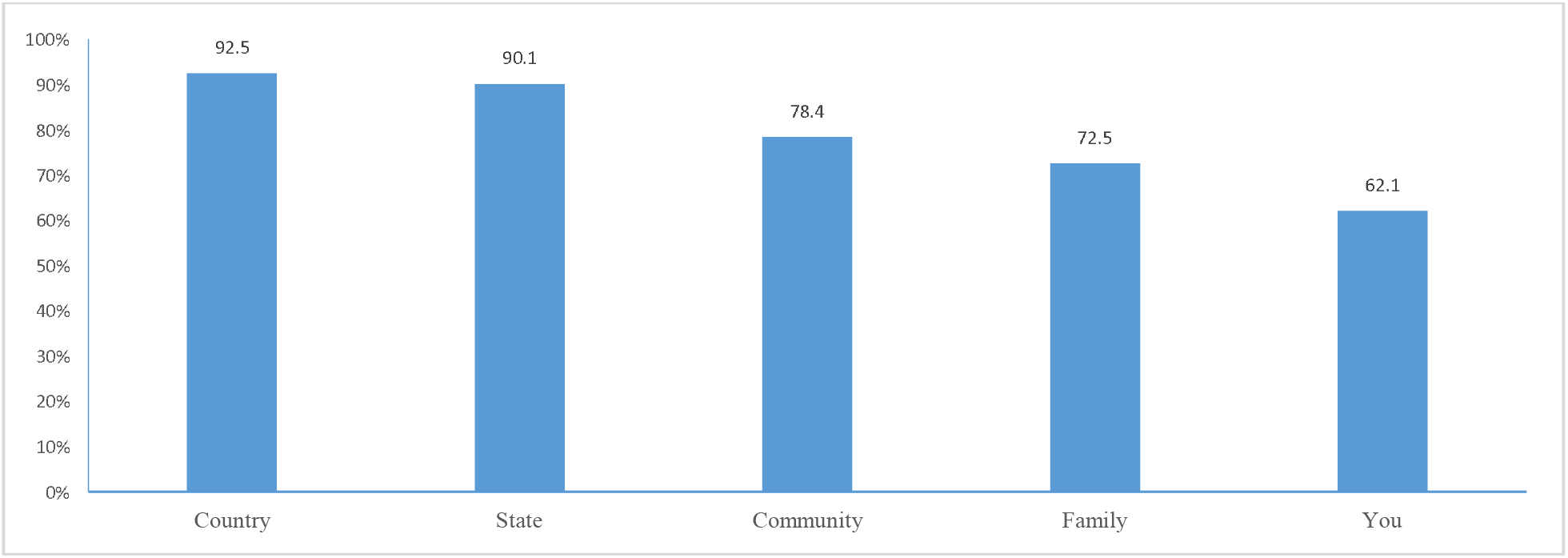
Perceived seriousness of COVID-19 for the country, the state, community, family and participants

### Knowledge about COVID-19

When asked about their knowledge of COVID-19 symptoms, most participants recognized cough (97.2%), shortness of breath (96.6%), fever (96.3%), fatigue (91.5%). Other responses were headache (54.7%) and diarrhea (37.5%) (Supplement Table B). However, respondents also incorrectly reported some symptoms that had not been identified and conveyed to the public and are not associated with COVID-19 including sneezing and pain in hands and feet.

### Adoption of preventive behaviors

In response to the question about what participants have done to reduce the chances of getting the coronavirus: nearly 100% reported frequently washing hands, avoiding contact with people (e.g. no hugs or handshakes) and avoiding events and meetings with a large number of people. Other behaviors included stayed at home as much as possible (98%), coughed and sneezed into the crook of the elbow (94.7%) and avoided public transportation (96.5%). Only 21.5% and 9.2% reported wearing sanitary gloves and wearing a mask when going outside respectively (Supplement Figure D).

When asked what they would do if they have symptoms COVID-19, the majority of the participants (92.8%) would call their primary care provider and stay in touch with the primary care provider if they felt they had any symptoms of COVID-19. Another frequent response was calling a hotline for guidance (71.0%). In order to protect family members, almost all indicated quarantining from other family members and visitors (98.5%) and staying away from work or other public places (99.6%) (Supplement Table C).

### Multivariate analysis of factors associated with anxiety, perceived seriousness of COVID-19 and loneliness during the social isolation

Results from logistic regression analysis (Table 2) documented that individuals with a higher level of COVID-19 knowledge were more likely to report a higher level with anxiety (OR:1.20, 95% CI: 1.03-1.41, p=0.022). No statistically significant differences were found in COVID-19 knowledge, self-rating seriousness of COVID-19 and loneliness. Those who stockpiled food and supplies for three to four weeks were more likely to have greater level of loneliness (OR=1.81, 95%CI:1.06-3.09, p=.030) than those who stockpiled two weeks or less. Being married was related to higher levels of anxiety about the virus (OR=1.79, 95% CI:1.07-2.99, p=0.026). Women were less likely to report loneliness than men (OR=0.37, 95% CI:0.19-0.69, p=0.026). Older age was associated with taking the pandemic more seriously (OR=1.03, 95% CI: 1.01-1.05, p=.025) and was also associated with loneliness during the social isolation (OR:1.05, 95% CI: 1.03-1.07, p<0.001)

**Table 2.** Multivariate logistic regression analysis of correlates of anxiety, perceived seriousness of COVID-19 and loneliness.

| Variables | Anxiety<sup>a</sup> |  | Perceived seriousness<sup>b</sup> |  | Loneliness<sup>c</sup> |  |
| --- | --- | --- | --- | --- | --- | --- |
|  | OR (95% CI) | P | OR (95% CI) | P | OR (95% CI) | P |
| **Age** | 0.99 (0.97-1.01) | 0.403 | 1.03 (1.01-1.05) | **0.025** | 1.05 (1.03-1.07) | **<0.001** |
| **Education** | 1.02 (0.97-1.08) | 0.387 | 1.02 (0.97-1.09) | 0.327 | 1.03 (0.97-1.09) | 0.346 |
| **Gender** |  |  |  |  |  |  |
| Male | 1 |  | 1 |  | 1 |  |
| Female | 2.18 (1.24-3.87) | **0.007** | 1.23 (0.68-2.21) | 0.485 | 0.37 (0.19-0.69) | **<0.001** |
| **Marital status** |  |  |  |  |  |  |
| Others<sup>d</sup> | 1 |  | 1 |  |  |  |
| Married or living with partners | 1.79 (1.07-2.99) | **0.026** | 2.21 (1.29-3.75) | **0.040** | 1.52 (0.87-2.68) | 0.140 |
| **Ethnicity** |  |  |  |  |  |  |
| Non-White<sup>e</sup> | 1 |  | 1 |  | 1 |  |
| White | 1.01 (0.62-1.66) | 0.967 | 1.25 (0.72-2.07) | 0.447 | 0.93 (0.55-1.59) | 0.794 |
| **Stockpile** |  |  |  |  |  |  |
| Two weeks or less | 1 |  | 1 |  | 1 |  |
| Three to four weeks | 1.05 (0.64-1.72) | 0.860 | 1.06 (0.62-1.81) | 0.845 | 1.81 (1.06-3.09) | **0.030** |
| **COVID-19 knowledge<sup>f</sup>** | 1.20 (1.03-1.41) | **0.022** | 1.03 (0.87-1.22) | 0.680 | 0.95 (0.81-1.23) | 0.591 |
| **Sources of information<sup>g</sup>** | 1.01 (0.85-1.21) | 0.911 | 1.01 (0.83-1.22) | 0.894 | 0.90 (0.75-1.00) | 0.277 |

Note. CI= confident interval; OR: odds ratio.
<sup>a,b</sup>The outcomes were categorized into a value of 1 corresponding to answers of “likely” or “very likely” and a value of 0 for all other responses to the question.
<sup>c</sup>Loneliness was categorized into a value of 1 corresponding to answers of “Yes” and a value of 0 for responses of “No”.
<sup>d</sup>Others including single, never married, divorced or separated and widowed.
<sup>e</sup>Non-White including African American, Latino, Asian and others.
<sup>f</sup>Correct answers of nine questions on COVID-19 knowledge were summed up to obtain overall score with higher scores indicating greater levels of COVID-19 knowledge;
<sup>g</sup>Total number of eight sources of information was summed up to obtain total score, with higher scores indicating greater number of sources of information.

## Discussion

To our knowledge, this is first assessment of public knowledge, perceived risk and resulting behavior changes in response to the COVID-19 outbreak in during an early phase of social distancing and isolation. This study provides real time and necessary information for the public health agencies and authorities to strategically respond to community needs and convey effective public health messages to the community experience the COVID-19 outbreak.

The participants in this survey were well-articulated to informational sources and had greater trust in scientifically based informational sources. There was good adherence with the preventive behavioral recommendations among participants at the time. They also had accurate knowledge consistent with the official messaging. While only few participants stated wearing sanitary gloves and facemasks when going outside, these messages were consistent with recommendations by the CDC at this time. Wearing facemasks is now mandatory when going to the public places in the state.

However, participants underestimated their own risk for the COVID-19 infection and its seriousness as the findings show that a significant number of respondents identified the virus as serious for the country, the state, and their community more than for their family and themselves. This finding provides further support for the well-documented phenomenon in the social psychological literature on coping with threatening events referred to as ‘unique invulnerability’ in which people view themselves as less vulnerable than others to serious outcomes/adversities.^13^ It could be that with the enormous flow of information, people see a grimmer picture of the pandemic at the broad level of country and state, but when people look at their family and friends, they still see few signs of impact. This phenomenon may contribute to less conformity to behavioral guidelines. Thus, conveying timely and effective information about the seriousness of the COVID-19 is crucial to ensure that people do not minimize the seriousness of the virus.

Interestingly, individuals with higher levels of COVID-19 knowledge were more likely to report a higher level of anxiety. Absorbing constant information and news about the virus might affect participants’ mental status and increase their levels of anxiety. Reducing information updates about the virus might help reduce the level of anxiety but also has the effect of not staying current with information about risk and prevention. It is noticeable that nearly half of the participants in this study experienced a high level of anxiety. Therefore, it is important for public health agencies and authorities not only provide information about COVID-19, but addressing mental health as well. It is crucial that messaging to the public requires not only status reports and behavioral guidelines, but also including a component of positive information that can reduce anxiety.

In this study, being married was more likely to increase their anxiety level. Perhaps married participants may be concerned about impact of COVID-19 not only about themselves, but also about their children and partners. Females reported higher level of anxiety about COVID-19 which is consistent with findings of a recent study among Iranian during COVID-19 outbreak.^14^ However, women were less likely to report loneliness than males, perhaps because women may get busier in times of crisis or they may have a greater social network than men, and thus, have less feeling of loneliness.

Respondents reported accessing a variety of sources of information on COVID-19. A large number of respondents received information through electronic media, family members and friends indicating that traditional forms of electronic media (e.g. TV) and communications through family and friend networks remain the primary information sources and channels for public health messages. Noticeably, over 80% of participants accessed information about COVID-19 via social media. Social media have increasingly become an important source of information and an inexpensive communication medium that allows health agencies to quickly disseminate their messages to potentially large audiences.^15^ The study findings suggest that public health and government agencies should take advantage of social media outlets to convey public health communications and information about COVID-19 to the population. However, it is essential that the information is endorsed and approved by technical agencies such as the Centers for Disease Control and Prevention (CDC) and the National Institutes of Health (NIH) before the information is posted. Social media can cause confusion if the information is not accurate and credible. A recent study in the U.S. and UK found that while participants generally had good knowledge of the main COVID-19 transmission modes and common symptoms, they reported misconceptions on COVID-19 prevention derived from social media, including discrimination against individuals of East Asian ethnicity because they might spread COVID-19.^11^

This rapid and “real time” assessment should be interpreted with caution in light of its limitations, which include a non-random convenience sample, which is not representative of residents in Connecticut. The nature of this cross-sectional survey does not allow to infer causality. This survey sample provided us a good assessment of a more educated and predominantly female subpopulation and the results will be used revise the survey questions for subsequent administration of surveys focusing on underrepresented and under-resourced subpopulations. It is also not geographically representative of residents in Connecticut. While this survey provides insights about a respondent’s reaction to the COVID-19, additional qualitative methods would allow for deeper insight, further informing effective public health communications.

### Public Health Implications

The COVID-19 pandemic has affected and taken a heavy toll on every aspect of respondents’ lives. In this study, while participants reported good knowledge and adherence with the preventive behavioral recommendations, a large number of participants experienced a high level of anxiety and loneliness during the isolation in the early phase of the COVID-19 pandemic. It is crucial for the public health authorities not only provide accurate and scientific information about the virus promoting protective behavior changes but also to minimize anxiety through supportive messages and recommendations for positive coping strategies. This might include timely offering counselling services for those in need right from the onset of the pandemic.

## Data Availability

Data was collected using Qualtrics software and is available in the Qualtrics

---

**Table A:** Source of information about COVID-19 (n=464)

| **Sources of information** | **No** | **%** |
| --- | --- | --- |
| Electronic media and TV | 390 | 85.5 |
| Social media | 369 | 81.3 |
| Printed media | 184 | 41.1 |
| Healthcare provider/hospital | 280 | 61.9 |
| Family members | 312 | 69.0 |
| Friends | 309 | 68.7 |
| Clergy | 38 | 8.2 |

**Table B:** Knowledge about COVID-19 symptoms (n=464)

| **Symptoms** | **No** | **%** |
| --- | --- | --- |
| Dry cough | 448 | 97.2 |
| Shortness of breath | 437 | 96.6 |
| Fever | 443 | 96.3 |
| Fatigue | 419 | 91.5 |
| Body aches | 337 | 73.7 |
| Headache | 250 | 54.7 |
| Diarrhea | 172 | 37.6 |
| Sneezing | 117 | 25.9 |
| Pain in hands and feet | 34 | 7.5 |

**Table C.** Health seeking behaviors if having symptoms of COVID-19 (n=464)

| **Options** | **No** | **%** |
| --- | --- | --- |
| Go immediately to the emergency room | 9 | 2.0 |
| Calling a provider before visiting | 427 | 92.8 |
| Stay in touch with a primary care provider | 434 | 94.8 |
| Call a hotline for guidance | 323 | 71.0 |
| Seek a COVID-19 test | 265 | 58.1 |
| Stay at home, rest and monitor symptoms | 441 | 96.7 |
| Quarantine myself from other family members and visitors | 453 | 98.5 |
| Stay away from work or other public places | 457 | 99.6 |

**Figure C.**
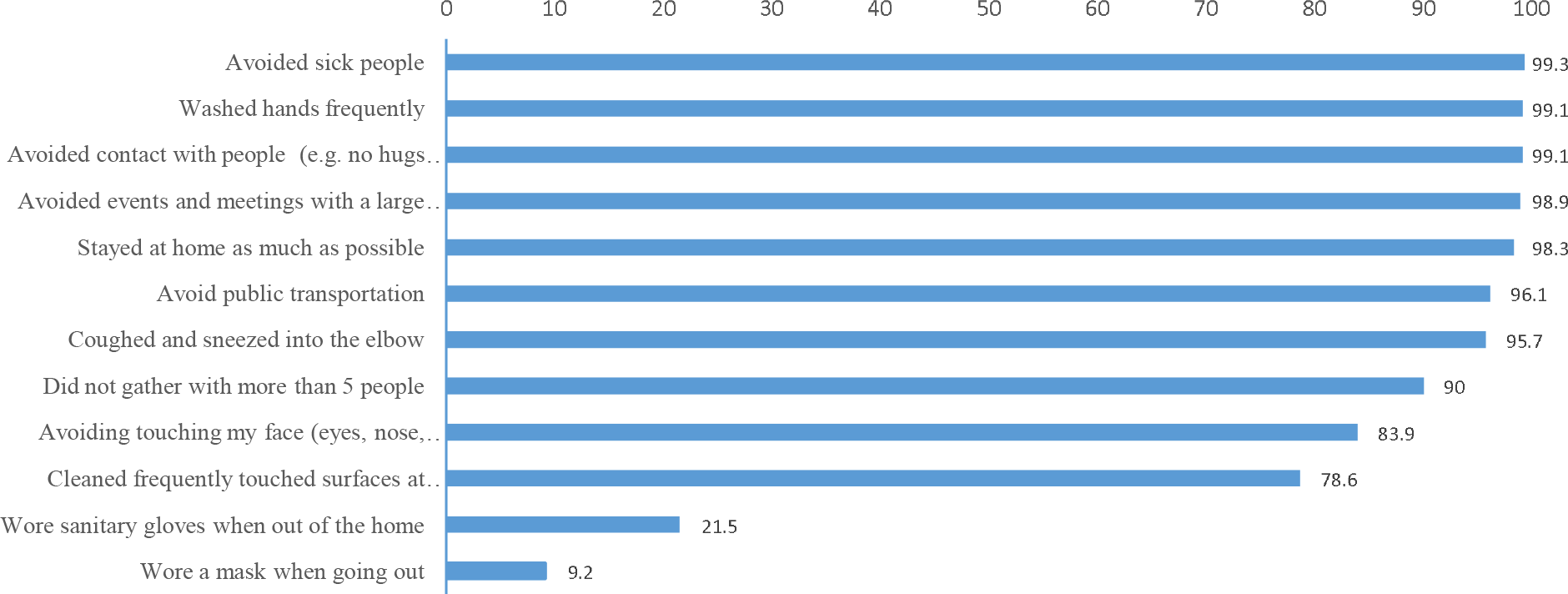
Adoption of preventive behaviors to COVID-19

## Notes

### Competing Interest Statement

The authors have declared no competing interest.

### Funding Statement

No funding support

